# Physiological pattern of cognitive aging

**DOI:** 10.1101/2021.08.04.21261599

**Authors:** Jan S. Novotný, Juan P. Gonzalez-Rivas, Maria Vassilaki, Janina Krell-Roesch, Yonas E. Geda, Gorazd B. Stokin

## Abstract

The effect of aging on cognition in cognitively healthy adult populations remains poorly investigated. Given that cognition evolves in time and thus, during aging, and becomes eroded in several disorders, some of which have recently acquired biological definitions, it is imperative to understand the process of cognitive aging. To determine the association of aging with cognitive performance in a cognitively healthy population, we studied cognitive performance in population-based cohort of 673 adults (aged 25-89). We found a gradual decline in cognitive performance across the lifespan, which requires two decades to demonstate significant change. This age-related decline was not significantly altered by either gender or education. These findings contribute to understand cognitive aging and provide essential data on physiological cognition for more precise diagnostics and timely intervention of early changes in cognition.

## Introduction

Although the effect of aging on cognitive performance is considered “common knowledge,” no recent study provides evidence supporting this statement. Knowing the pattern of physiological cognitive aging is critical for several reasons. Populations across the world’s are aging^1^, people in both, high and low-income counties are getting older and often work and maintain more active lifestyles until later in life. Despite the fact that previous studies occasionally reported specific aspects of cognitive aging^2^, there are no recent studies systematically measuring the effects of aging on cognition in modern population-based samples, characterized by unprecedented longevity.. Last, old age is the major risk factor for Alzheimer’s and related disorders and underlying pathological changes occur several years prior to the onset of the clinical symptoms, therefore, when individuals are cognitively unimpaired. Therefore, understanding the process of cognitive aging is critical for the design and conduct of effective interventions, diagnostics and therapy. In this study, we examined the physiological cognitive performance during aging in a well-characterized adult population-based sample.

## Results

We identified 848 out of 2434 participants enrolled in the Kardiovize study that underwent cognitive testing. 149 were excluded due to missing more than 10% of the demographic data or incomplete cognitive testing results. 699 MoCA total scores split into the age groups are presented in the supplemental Table 1 (eTable 1). 26 participants were excluded due to MoCA scores indicative of MCI (eFigure 1). The final sample consisted in 673 cognittively unimpaired participants. Of these participants, 353 (52%) were females,and the mean age was 52.3 ±14.2 (Table 1).

**Table 1.**
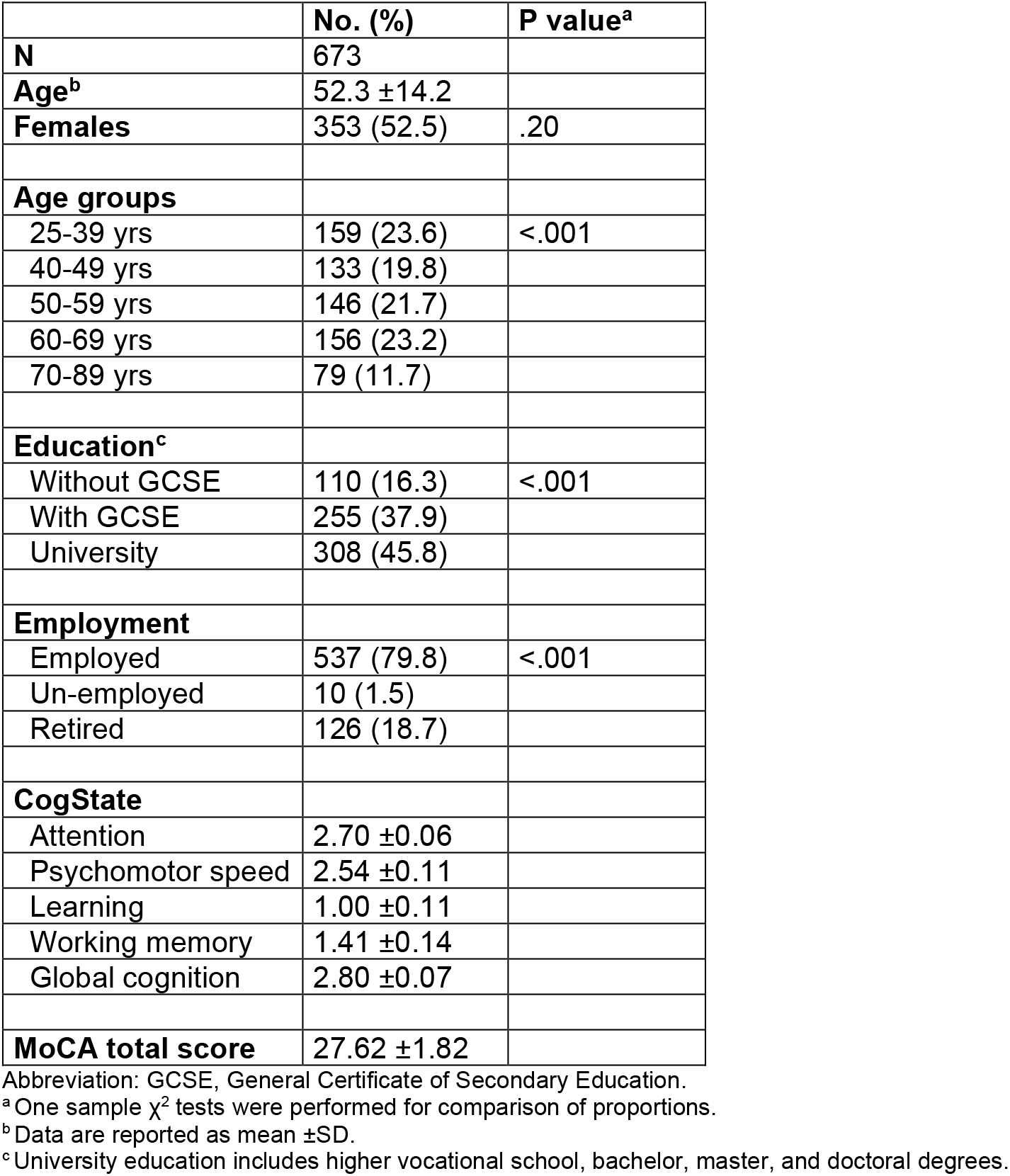
Basic characteristics of the sample.

|  | No. (%) | P value<sup>a</sup> |
| --- | --- | --- |
| **N** | 673 |  |
| **Age<sup>b</sup>** | 52.3 ±14.2 |  |
| **Females** | 353 (52.5) | .20 |
| **Age groups** |  |  |
| 25-39 yrs | 159 (23.6) | <.001 |
| 40-49 yrs | 133 (19.8) |  |
| 50-59 yrs | 146 (21.7) |  |
| 60-69 yrs | 156 (23.2) |  |
| 70-89 yrs | 79 (11.7) |  |
| **Education<sup>c</sup>** |  |  |
| Without GCSE | 110 (16.3) | <.001 |
| With GCSE | 255 (37.9) |  |
| University | 308 (45.8) |  |
| **Employment** |  |  |
| Employed | 537 (79.8) | <.001 |
| Un-employed | 10 (1.5) |  |
| Retired | 126 (18.7) |  |
| **CogState** |  |  |
| Attention | 2.70 ±0.06 |  |
| Psychomotor speed | 2.54 ±0.11 |  |
| Learning | 1.00 ±0.11 |  |
| Working memory | 1.41 ±0.14 |  |
| Global cognition | 2.80 ±0.07 |  |
| **MoCA total score** | 27.62 ±1.82 |  |

Abbreviation: GCSE, General Certificate of Secondary Education.
<sup>a</sup> One sample $\chi^2$ tests were performed for comparison of proportions.
<sup>b</sup> Data are reported as mean ±SD.
<sup>c</sup> University education includes higher vocational school, bachelor, master, and doctoral degrees.

Mean cognitive performance (mean [SD]) of the cohort was 2.70 [0.06], 2.54 [0.11], 1.00 [0.11], 1.41 [0.14], and 2.80 [0.07] for attention, psychomotor speed, learning, working memory, and global cognition, respectively (Table 1). Participants had better cognitive performance in psychomotor speed compared to attention, and in working memory compared to learning. In relation to aging, the cognitive scores declined significantly from 2.67 [0.05] to 2.72 [0.05] for attention, from 2.49 [0.07] to 2.61 [0.10] for psychomotor speed, from 1.04 [0.10] to 0.97 [0.10] for learning, from 1.45 [0.12] to 1.35 [0.18] for working memory, and from 2.76 [0.05] to 2.84 [0.07] for global cognition in the oldest to the youngest age groups (Table 2, Figure 1).

**Table 2.**
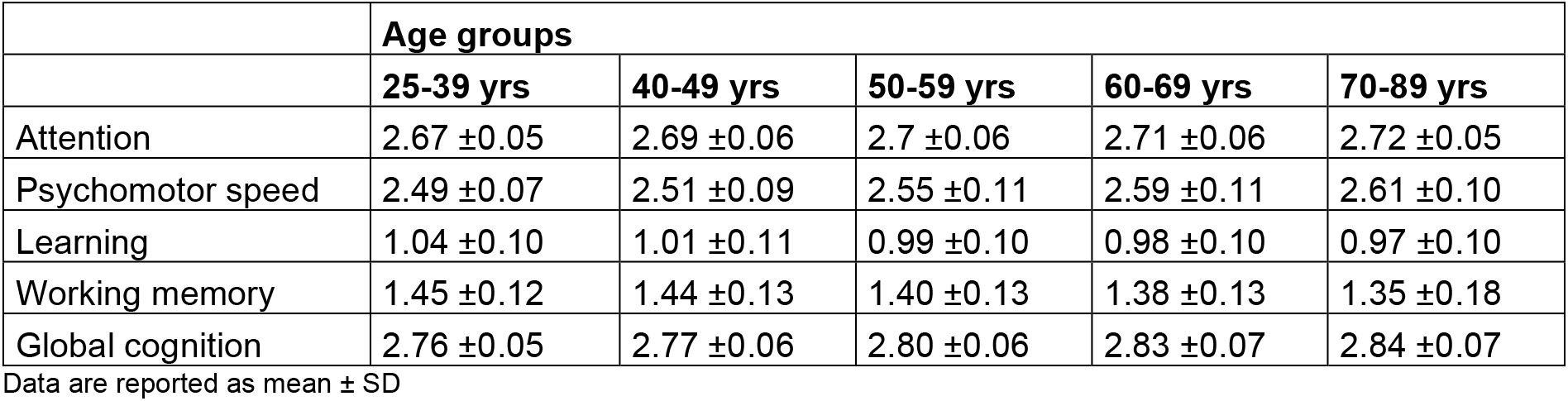
Characteristics of cognitive ageing in individual cognitive domains.

**Figure 1.**
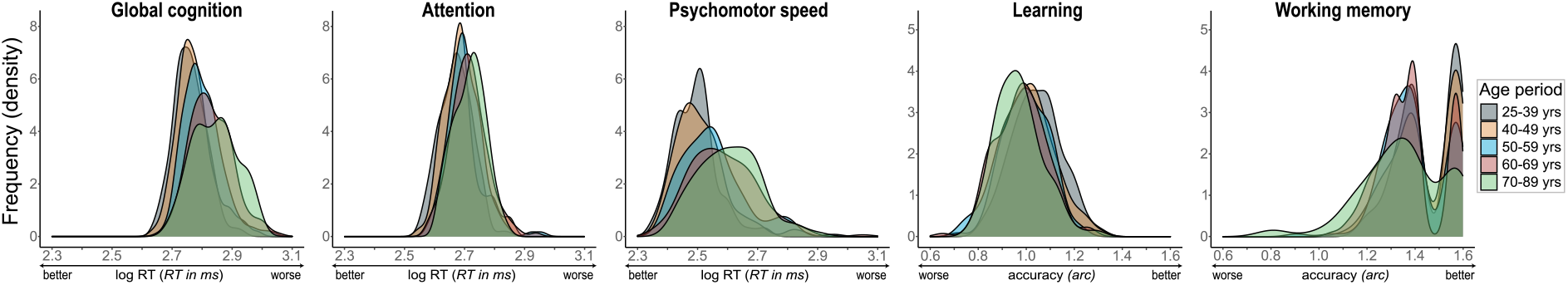
Distribution of CogState variables in individual age groups. Frequency (y) was calculated using probability density function based on Kernel density estimation.

ANOVA and ANCOVA models confirmed a significant age-related difference in the performance in all cognitive domains. Age proved to be the most significant factor, as neither sex nor education substantially altered the significance and the effect size of the models (Table 3). The largest aging effect was observed in psychomotor speed (P<.001, part. η^2^=0.112, medium effect) and global cognition (P<.001, part. η^2^=0.144, large effect). Differences in cognitive performance measurements required two decades for a significance to be observed (eTable 2).

**Table 3.** ANOVA/ANCOVA models: effect of age on cognitive performance.

|  | **Model 1: Effect of age<sup>a</sup>** |  |  | **Model 2: Effect of age adjusted for sex<sup>b</sup>** |  |  | **Model 3: Effect of age adjusted for education<sup>b</sup>** |  |  | **Model 4: Effect of age adjusted for sex and education<sup>b</sup>** |  |  |
| --- | --- | --- | --- | --- | --- | --- | --- | --- | --- | --- | --- | --- |
|  | **F** | **P** | **part. $\eta^2$** | **F** | **P** | **part. $\eta^2$** | **F** | **P** | **part. $\eta^2$** | **F** | **P** | **part. $\eta^2$** |
| Attention | 15.047 | <.001 | 0.083 | 13.593 | <.001 | 0.075 | 11.171 | <.001 | 0.063 | 10.093 | <.001 | 0.057 |
| Psychomotor speed | 31.100 | <.001 | 0.157 | 27.551 | <.001 | 0.142 | 23.585 | <.001 | 0.124 | 20.926 | <.001 | 0.112 |
| Learning | 11.394 | <.001 | 0.064 | 10.754 | <.001 | 0.061 | 7.598 | <.001 | 0.044 | 7.310 | <.001 | 0.042 |
| Working memory | 11.078 | <.001 | 0.062 | 11.422 | <.001 | 0.064 | 7.498 | <.001 | 0.043 | 7.850 | <.001 | 0.045 |
| Global cognition | 39.144 | <.001 | 0.190 | 32.247 | <.001 | 0.174 | 30.781 | <.001 | 0.156 | 27.827 | <.001 | 0.144 |

Abbreviation: part. $\eta^2$ , partial Eta-squared.
<sup>a</sup> ANOVA tests were performed for comparison of means.
<sup>b</sup> General Linear Models (ANCOVA type) were performed for comparison of means while adjusting for the effect of sex and education.

Age-related changes in cognitive performance were manifested, in addition to a decline in the mean scores, also by significantly increased variance in values at older ages. This was reflected both in larger standard deviations and in the significantly increased coefficient of variation. While attention and learning maintained stable variation throughout aging, global cognition showed a steady slow increase, psychomotor speed variance first increased with age and plateau-ed out in older age, while working memory continued increasing throughout the lifespan with a distinct rise in the oldest age (global cognition, CV=1.93, 2.21, 2.23, 2.48, 2.61, P=.011, psychomotor speed, CV=2.86, 3.53, 4.23, 4.36, 3.91, P<.001, and working memory, CV=8.16, 9.02, 9.14, 9.44, 12.96, P<.001, for age of 25-39, 40-49, 50-59, 60-69, 70-89, respectively) (Figure 2, eTable 3).

**Figure 2.**
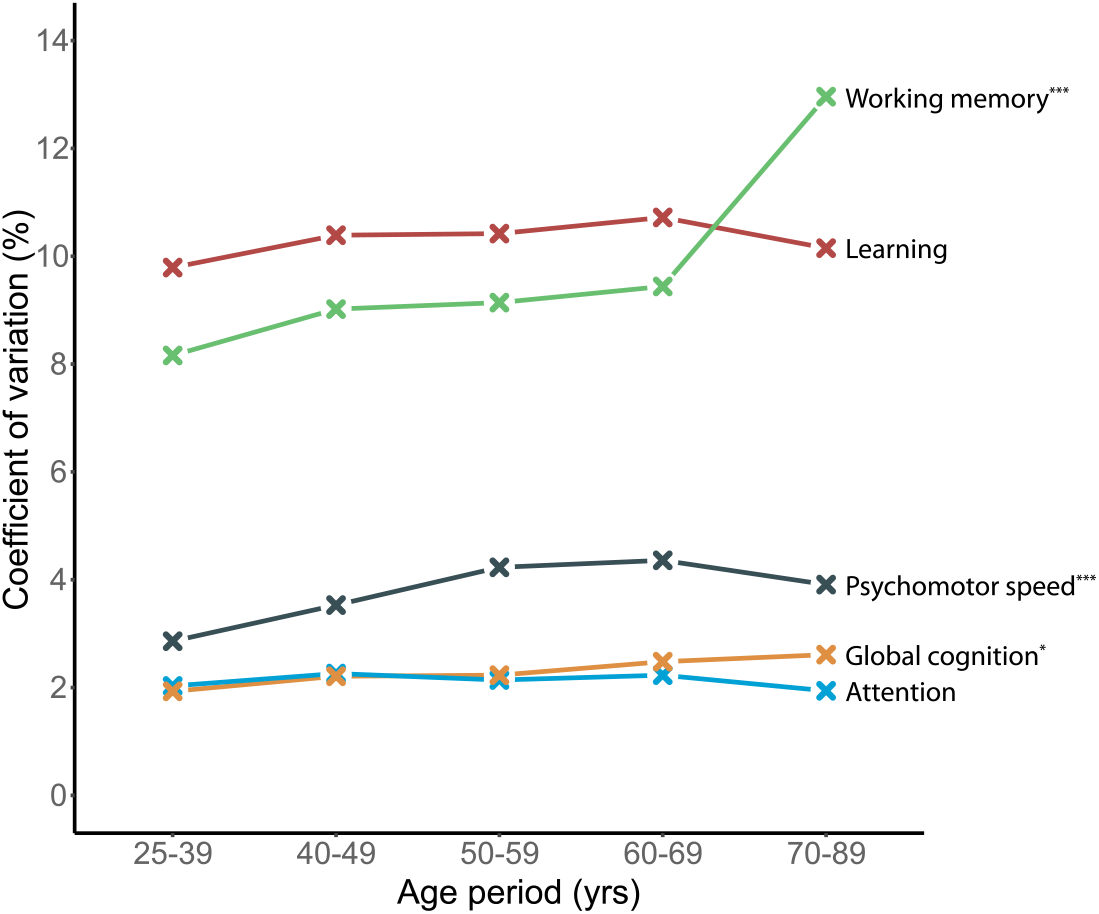
Coefficients of variation of CogState variables in individual age groups. The *P* values were obtained from Signed-Likelihood Ratio Test.

## Discussion

We observed an age-related progressive decline in cognitive performance across the lifespan. Declined cognitive performance in the oldest age groups was detected in global and domain-specific cognition. Our measurements showed that cognitive performance significantly decreased in two decades and this difference was observed throughout the adulthood. These findings indicate that cognitive decline is a relatively slow process in a cognitively healthy population, beginning as early as in midlife. In parallel to the differences in mean cognitive performance, the variability in cognitive performance also increased with age, suggesting greater interindividual differences in cognition at older ages. Most of the domains were more prone to gradual lower mean values through the lifespan and plateau-ed out at older age with attention and learning showing more compact distribution compared to the others. This suggests that physiological changes in cognition later in life are likely to be more intricately linked to confounding factors including risk factors. Working memory showed a different pattern, which was characterized both, by significantly higher variability and a significant jump in interindividual differences at older ages.

To the best of our knowledge, there are no studies in the last decades describing cognitive performance in different age groups in a cognitively healthy population. In fact, most studies focus on different issues such as longitudinal changes in cognition after injury or in other disorders or in relation to risk factors^10,11^. As a result, determination of healthy cognitive performance in relation to aging is merely a by-product of these studies. Our findings measure and formalize the process of cognitive aging in a well-characterized cognitively healthy population, which is of relevance due to several reasons.

First, considering the aging of today’s populations^1^, by knowing physiological cognitive aging, it might be possible to detect in a timely manner new age-related changes of cognition that might be pathological and/or linked to other disorders that previously were not known. Second, current diagnostics are often based on the detection of deficits by using predefined cut-off points. This approach has some shortcomings in that the cut-off points may not be accurate or may change over time^12–15^, and are often based on the overall score of the instrument, thus failing to capture changes in individual cognitive domains. Comparing individual to typical age-specific cognitive profiles in addition to screening for cognitive deficits may well improve early detection of cognitive disorders. Furthermore, cognitive profiles offer more detailed insight into changes in individual domains and their interactions. Finally, as biological definitions of cognitive disorders were recently reported, and a new medication for Alzheimer’s disease was just recently approved for clinical treatment after almost 20 years^16^, there is a great need for a precise and timely identification of early changes in cognition. Without knowledge of the pattern of physiological cognitive performance across the lifespan, such identification and evaluation are difficult.

First, our study sample, although generally healthy and well-characterized, was relatively highly educated compared to the general education level in the population, which might have affected the observed overall cognitive patterns^17,18^. This might be linked to the city-based nature of the study sample, and inclusion of more variable population might have led to slightly different results. However, the models adjusted for education show stable unchanged age-related differences in cognition. Second, given the generally low prevalence of diseases in the study population, it is possible that this good level of health positively affected the cognitive performance of the older group compared to similarly aged non-participants with a higher prevalence of diseases. Finally, although we rigorously tested the cognitive performance with adjusting for key possible confounders, the cross-sectional nature of the data does not allow for causal inferences and longitudinal conclusions to be made. We formalized characteristics of global and domain-specific cognitive differences between the age groups globally and described the connection between cognition and aging. The results showed gradually deteriorating cognitive performance across the lifespan with two decades needed for significant change to be observed. These findings have various implications. First, they may contribute to understanding the process of cognitive aging. Second, they might help to develop more precise diagnostics of current and new disorders using a cognitive profile-based approach. And last, they might lead to a more accurate and timely intervention, especially with regard to domains such as working memory, which showed great variability with age and may be amenable to targeted interventions.

## Methods

### Participants and Study Design

The research was conducted in the setting of the Kardiovize study, a longitudinal epidemiological cohort based on a randomly selected 1% population sample of the residents of the city of Brno, Czech Republic^3^. All study participants with data on cognitive performance were included in the study. To test for age-related changes, we categorized participants into 25-39, 40-49, 50-59, 60-69, and 70-89 year old age groups. Data were entered into a validated web-based research electronic data capture (REDCap) database^4^. The research protocols of the study were approved by the Institutional Review Board and by the ethics committee of St. Anne’s University Hospital, Brno, Czech Republic. All participants of the Kardiovize study provided written informed consent.

### Measurements

The Montreal Cognitive Assessment (MoCA) was used to identify subjects with pathological cognitive performance such as mild cognitive impairment (MCI) and dementia^5^. The MoCA total score ranges from 0 to 30, with a higher score indicating better cognitive performance. A MoCA score of < 23 was considered indicative of MCI^6^.

Physiological cognitive performance was assessed using the well-established Cogstate® Brief Battery (CBB). CBB is a short version of the computer-administered cognitive test battery requiring roughly 10 minutes for administration^7–9^. It uses playing cards to examine four basic cognitive domains: visual attention, psychomotor speed, visual learning, and working memory. Attention and psychomotor speed were assessed by measuring the response time needed to correctly identify the red playing cards (identification) or to detect all new playing cards (detection), respectively. The primary outcome measure of the attention and psychomotor speed tasks was the log10 transformed reaction time of correct responses in milliseconds (*log RT (RT in ms)*). Learning and working memory were assessed by measuring accuracy in recognizing a card previously seen in the deck (one card learning) or determining whether the shown card is the same as the last one (one back test), respectively. The primary outcome measured was the arcsine of the square root of the correct responses (*arc*). Mean *log RT (RT in ms)* of all four cognitive domains were used to calculate the global cognition score.

### Statistical analysis

Descriptive statistics were performed on demographic variables. No missing values were detected. Demographic data were compared between sexes, age groups, and education using one sample χ^2^ test for categorical variables. ANOVA and ANCOVA tests and Games-Howell post-hoc tests were performed to assess age-related differences in cognitive performance. Overall and pair differences in coefficients of variation were examined using the Signed-Likelihood Ratio Test.

Any 2-sided *P*<.05 was considered statistically significant. Statistical analyses and data visualizations were performed using SPSS (version 21) and R (v.3.6.3) using the ggplot2 (v.1.0.12) package.

## Supporting information

eTable

## Data Availability

The data used in this study are available on request immediately following the publication to anyone who submits the online request that will be approved by the St. Anne's University Hospital International Clinical Research Centre internal board. The researches have to provide their research intentions and goals, and specify, and justify requested variables. The data will be provided for a limited and well-defined time via cloud service or e-mail in csv format. After defined period the data should be returned, and all other copies should be destroyed.

## Author Contributions

GBS and JSN had full access to all the data in the study and take responsibility for the integrity of the data and the accuracy of the data analysis. The authors contributed to this article as follows: GBS conceived the idea of this study, JSN made the statistical analysis, JSN and GBS wrote the draft of the manuscript, JPGR, MV, JKR, YEG contributed to the critical revision of the manuscript for important intellectual content and reviewed.

## References

1. United Nations Department of Economic and Social Affairs, P. D. World Population Ageing 2020 Highlights: Living arrangements of older persons (ST/ESA/SER.A/451). (United Nations Publication, 2020).

2. Murman, D. The Impact of Age on Cognition. Semin. Hear. 36, 111–121 (2015).

3. Movsisyan, N. K. et al. Kardiovize Brno 2030, a prospective cardiovascular health study in Central Europe: Methods, baseline findings and future directions. Eur. J. Prev. Cardiol. 25, 54–64 (2018).

4. Harris, P. A. et al. Research electronic data capture (REDCap)—A metadata-driven methodology and workflow process for providing translational research informatics support. J. Biomed. Inform. 42, 377–381 (2009).

5. Nasreddine, Z. S. et al. The Montreal Cognitive Assessment, MoCA: A brief screening tool for mild cognitive impairment. J. Am. Geriatr. Soc. 53, 695–699 (2005).

6. Bartoš, A., Orlíková, H., Raisová, M. & Řípová, O. Czech Training Version of the Montreal Cognitive Assessment (MoCA-CZ1) for Early Identification of Alzheimer Disease. Ces. Slov Neurol N 77/110, 587–595 (2014).

7. Fredrickson, J. et al. Evaluation of the usability of a brief computerized cognitive screening test in older people for epidemiological studies. Neuroepidemiology 34, 65–75 (2010).

8. Darby, D. G. et al. Intraindividual cognitive decline using a brief computerized cognitive screening test. Alzheimer’s Dement. 8, 95–104 (2012).

9. Maruff, P. et al. Clinical utility of the cogstate brief battery in identifying cognitive impairment in mild cognitive impairment and Alzheimer’s disease. BMC Psychol. 1, (2013).

10. Baker, J. E. et al. Cognitive impairment and decline in cognitively normal older adults with high amyloid-β: A meta-analysis. Alzheimer’s Dement. Diagnosis, Assess. Dis. Monit. 6, 108–121 (2017).

11. Saa, J. P. et al. Longitudinal evaluation of cognition after stroke – A systematic scoping review. PLoS One 14, e0221735 (2019).

12. Elkana, O., Tal, N., Oren, N., Soffer, S. & Ash, E. L. Is the Cutoff of the MoCA too High? Longitudinal Data From Highly Educated Older Adults. J. Geriatr. Psychiatry Neurol. 33, 155–160 (2020).

13. Morgado, J., Rocha, C. S., Maruta, C., Guerreiro, M. & Martins, I. P. Cut-off scores in MMSE: a moving target? Eur. J. Neurol. 17, 692–695 (2010).

14. Gloster, A. T. et al. Impact of COVID-19 pandemic on mental health: An international study. PLoS One 15, e0244809 (2020).

15. Carson, N., Leach, L. & Murphy, K. J. A re-examination of Montreal Cognitive Assessment (MoCA) cutoff scores. Int. J. Geriatr. Psychiatry 33, 379–388 (2018).

16. FDA Grants Accelerated Approval for Alzheimer’s Drug | FDA. https://www.fda.gov/news-events/press-announcements/fda-grants-accelerated-approval-alzheimers-drug.

17. Guerra-Carrillo, B., Katovich, K. & Bunge, S. A. Does higher education hone cognitive functioning and learning efficacy? Findings from a large and diverse sample. PLoS One 12, e0182276 (2017).

18. Schneeweis, N., Skirbekk, V. & Winter-Ebmer, R. Does Education Improve Cognitive Performance Four Decades After School Completion? Demography 51, 619–643 (2014).

