## Supplementary material for "Physiological pattern of cognitive aging": eTable

### Supplementary materials for „Physiological pattern of cognitive aging“

**eTable 1. MoCA total scores in individual age periods**

| | mean ±SD |
|---|---|
| 25-39 yrs | 28.30 ±1.49 |
| 40-49 yrs | 28.29 ±1.48 |
| 50-59 yrs | 27.88 ±1.45 |
| 60-69 yrs | 26.92 ±1.85 |
| 70-89 yrs | 26.05 ±2.04 |

**eFigure 1. MoCA total score distribution in individual age periods.**

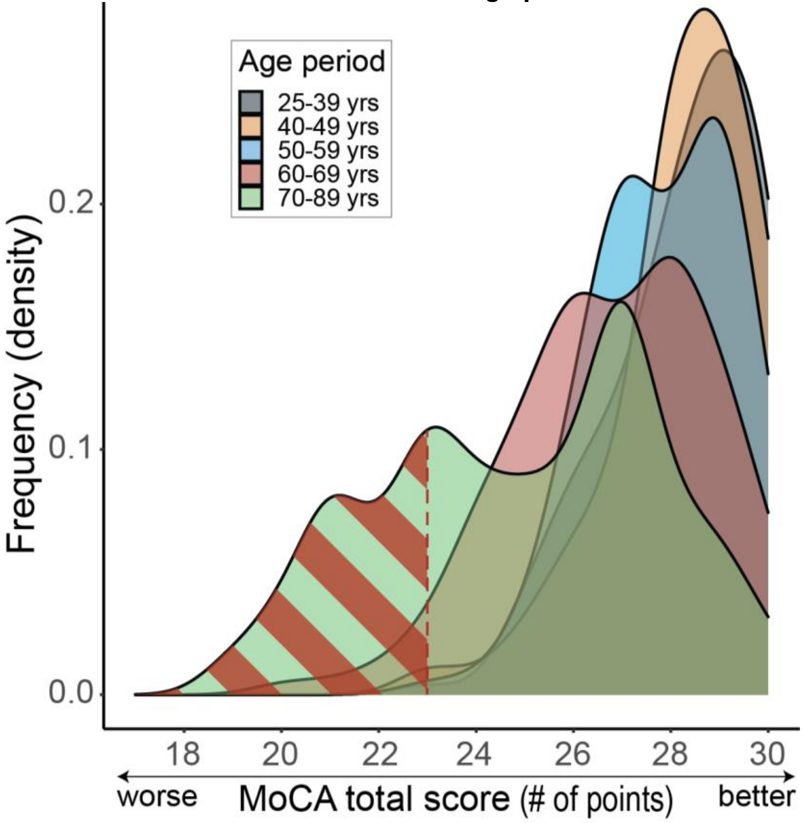


Red hatched area depicts scores consistent with MCI (<23). Frequency (y) was calculated using probability density function based on Kernel density estimation.

**eTable 2. Paired comparison (P-values) of between-group differences (Games-Howell post-hoc test) in cognitive performance between individual age periods**

| | | Attention | Psychomotor speed | Learning | Working memory | Global cognition |
|---|---|---|---|---|---|---|
| **25-39** | **40-49** | .32 | .27 | .12 | .94 | .23 |
| | **50-59** | <.001 | <.001 | <.001 | .005 | <.001 |
| | **60-69** | <.001 | <.001 | <.001 | <.001 | <.001 |
| | **70-89** | <.001 | <.001 | <.001 | <.001 | <.001 |
| **40-49** | **50-59** | .07 | .006 | .25 | .10 | .005 |
| | **60-69** | .001 | <.001 | .04 | .001 | <.001 |
| | **70-89** | .001 | <.001 | .02 | .002 | <.001 |
| **50-59** | **60-69** | .54 | .04 | .92 | .64 | .001 |
| | **70-89** | .43 | .002 | .64 | .20 | <.001 |
| **60-69** | **70-89** | >.99 | .72 | .96 | .73 | .63 |

**eTable 3. Paired comparisons (P-values) of between-group differences in coefficients of variation between individual age periods**

| | | Psychomotor speed | Working memory | Global cognition |
|---|---|---|---|---|
| **25-39** | **40-49** | .01 | .24 | .10 |
| | **50-59** | <.001 | .17 | .07 |
| | **60-69** | <.001 | .07 | .002 |
| | **70-89** | .001 | <.001 | .002 |
| **40-49** | **50-59** | .03 | >.99 | >.99 |
| | **60-69** | .01 | .25 | .18 |
| | **70-89** | .33 | <.001 | .10 |
| **50-59** | **60-69** | .77 | .75 | .21 |
| | **70-89** | .44 | <.001 | .12 |
| **60-69** | **70-89** | .27 | .001 | .65 |
